# The fourth dose of mRNA COVID-19 vaccine following 12 different three-dose regimens: Safety and immunogenicity to Omicron BA.4/BA.5

**DOI:** 10.1101/2023.01.19.23284761

**Authors:** Sitthichai Kanokudom, Jira Chansaenroj, Nungruthai Suntronwong, Suvichada Assawakosri, Ritthideach Yorsaeng, Pornjarim Nilyanimit, Ratchadawan Aeemjinda, Nongkanok Khanarat, Preeyaporn Vichaiwattana, Sirapa Klinfueng, Thanunrat Thongmee, Donchida Srimuan, Thaksaporn Thatsanatorn, Natthinee Sudhinaraset, Nasamon Wanlapakorn, Sittisak Honsawek, Yong Poovorawan

## Abstract

**Objective:** To investigate the reactogenicity and immunogenicity of the fourth dose using mRNA vaccines after different three-dose regimens and to compare the 30 µg BNT162b2 and 50 µg mRNA-1273 vaccines.

**Methods:** This prospective cohort study was conducted between June and October 2022. The self-recorded reactogenicity was evaluated on the subsequent 7 days after a fourth dose. Binding and neutralizing activity of antibodies against the Omicron BA.4/5 variants were determined.

**Results:** Overall, 292 healthy adults were enrolled and received BNT162b2 or mRNA-1273. Reactogenicity was mild to moderate and well-tolerated after a few days. Sixty-five individuals were excluded. Thus, 227 eligible individuals received a fourth booster dose of BNT162b2 (n=109) and mRNA-1273 (n=118). Most participants, regardless the type of previous three dose regimens, elicited a significantly high level of binding antibodies and neutralizing activity against the Omicron BA.4/5 28 days after a fourth dose. The neutralizing activity against the Omicron BA.4/5 between the BNT162b2 (82.8%) and mRNA-1273 (84.2%) groups was comparable with a median ratio of 1.02.

**Conclusion:** This study found that the BNT162b2 and mRNA-1273 vaccines can be used as a fourth booster dose for individuals who were previously immunized with any prior mix and match three dose COVID-19 vaccine.

## 1. Introduction

Coronavirus disease 2019 (COVID-19) is a flu-like disease caused by severe acute respiratory syndrome coronavirus 2 (SARS-CoV-2). SARS-CoV-2 has rapidly evolved across the world. Since the first identification of the Omicron variant in South Africa on 26 November 2021, several subvariants have been identified (Shrestha et al., 2022). The Omicron lineages have caused outbreaks of COVID-19 in multiple countries around the world from early 2022 to date (Tegally et al., 2022). On 6 January 2023, there were more than 667 million confirmed cases worldwide with 4.7 million cases in Thailand, and 6.7 million reported deaths worldwide and over 33,000 deaths in Thailand alone (Worldmeter, 2023).

In many countries, including Thailand, most initial vaccinations were administered with two doses of inactivated vaccine (CoronaVac or BBIBP-CorV), two doses of viral vector vaccine (AZD1222) or a heterologous schedule (CoronaVac followed by AZD1222) to ensure adequate coverage. After the arrival of BNT162b2 and mRNA-1273 in Thailand and the spread of the Delta variant SARS-CoV-2 in July 2021, the Ministry of Public Health recommended a third dose vaccination campaign with AZD1222, BNT162b2, and mRNA-1273 administered after three months of second dose vaccination. Subsequently, a fourth dose was offered on 21 March 2022, to combat the newly emerging Omicron variant, which evades pre-existing vaccine-induced immunity (Magen et al., 2022). On 2 December 2022, more than 26 million individuals (38.5%) of the Thai population had received three doses and approximately 5.8 million (8.4%) had received four doses (Thai-DDC, 2023). Limited safety and immunogenicity studies are currently available evaluating the fourth dose vaccination after three-dose mix-and-match vaccinations.

The third booster dose elicits a high immune response against infection (Akaishi et al., 2022; Wan et al., 2022). A field study demonstrated that the third dose increased vaccine effectiveness against disease severity (Accorsi et al., 2022, Favresse et al., 2022), and reduced hospitalization rates (Adams et al., 2022, Wan et al., 2022). Furthermore, a third dose vaccine increases neutralizing capacity against the Omicron variant in laboratory tests (Assawakosri et al., 2022a, Assawakosri et al., 2022b, Kanokudom et al., 2022a, Kanokudom et al., 2022b, Suntronwong et al., 2022b). Waning immunity after three doses appears to significantly reduce vaccine efficacy (VE) against infection at 3–6 months of follow-up (Favresse et al., 2022), and probably leads to breakthrough infections with the Delta/Omicron variants (Amanatidou et al., 2022). Recent studies have shown that a fourth dose of mRNA vaccine was associated with a reduced rate of symptomatic infection during the Omicron predominant period (Bar-On et al., 2022; Regev-Yochay et al., 2022). According to a cohort study of individuals aged 80 and older in Sweden, a fourth mRNA booster dose increased VE against mortality during the Omicron era compared to three doses of BNT162b2 (Nordström et al., 2022). Additionally, a previous report showed that the fourth dose reduced the infection rate and severity (Cohen et al., 2022).

Furthermore, surveillance of a total of 36,170 subjects in Northern Thailand indicated that VE against Omicron infection of the subjects receiving a fourth dose was 75%, followed by the three doses (VE of 31%), and one or two doses conferred little to no protection, respectively) (Intawong et al., 2022). However, the clinical outcomes of a fourth booster dose after different three-dose regimes remain unclear.

Many individuals require a booster dose to restore binding and neutralizing antibodies that are no longer sufficient to combat the emerging Omicron subvariants. This study investigated the increased effects of the fourth dose of mRNA vaccines, BNT162b2 and mRNA- 1273, after three mix and match dose regimens. In addition, we compared the reactogenicity and immunogenicity among these two booster groups. This study provides important data to help guide policy makers on the use of fourth doses of COVID-19 vaccines among the adult population in Thailand.

## 2. Materials and methods

### 2.1 Study designs and participant enrolment

The cohort study was conducted between June and October 2022 at the Center for Excellence in Clinical Virology, Chulalongkorn University, Bangkok. The study protocol was approved by the Institutional Review Board (IRB) of the Faculty of Medicine of Chulalongkorn University (IRB 223/65) and was conducted in accordance with the principles of the Declaration of Helsinki. This trial was registered in the Thai Registry of Clinical Trials (TCTR 20210910002). Written informed consent was obtained from the participants before enrollment.

A total of 292 healthy adults 18 years and older, who had previously been immunized with any of the three available doses of the COVID-19 vaccine for at least 3 months, were enrolled. The cohort was stratified into 12 groups according to their primary three doses (**Figure 1**). Participants with no history of COVID-19 disease received a fourth booster dose of 30 μg BNT162b2 or 50 μg mRNA-1273. Exclusion criteria were characteristics associated with serious medical conditions and individuals who had pre-existing immunoglobulin (Ig) G specific to nucleoprotein (N) (anti-N IgG), anti-N IgG seroconversion (negative to positive) and confirmed positive tests to SARS-CoV-2.

**Figure 1.**
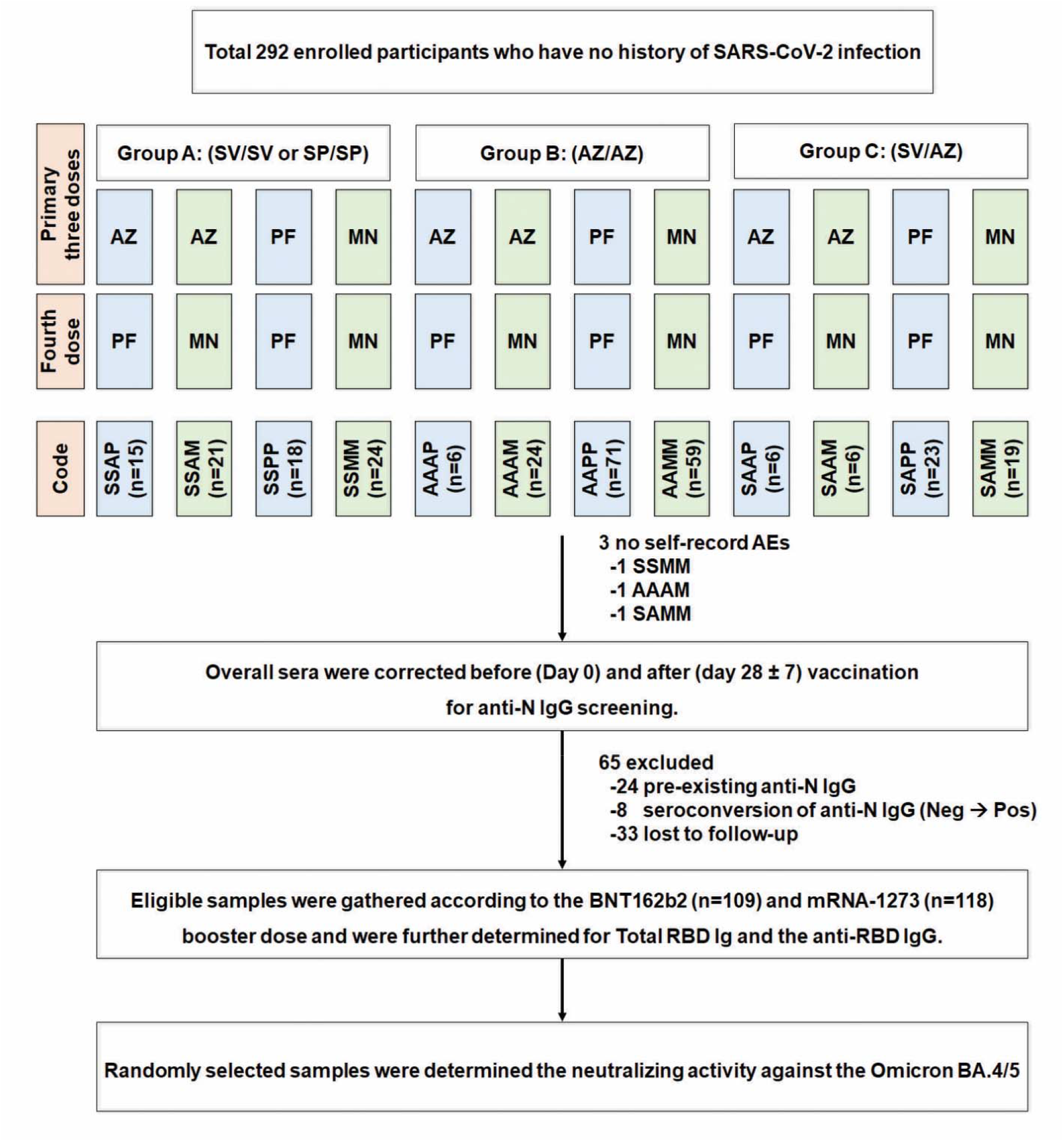
Study flow diagram of participants enrolled and collection of eligible samples. Individuals (18 years and older), who had previously been immunized with different three doses, were assigned to receive the fourth booster dose with 30 µg BNT162b2 or 50 µg mRNA-1273. Blood samples were collected on days 0 and 28 ± 7 after booster dose. Serum samples were previously screened according to the level of anti-nucleoprotein (N) IgG. Participants who had seropositive anti-N IgG (pre-existing and seroconversion) were excluded. The remaining participants were eligible for the evaluation of immunogenicity analyses after booster vaccination. Abbreviations: AEs, adverse events; A and AZ, AZD1222; Ig; immunoglobulin; M and MN, mRNA-1273; N, nucleoprotein; P and PF, BNT162b2; S, inactivated vaccine BBIBP-CorV (SP) or CoronaVac (SV).

### 2.2 Vaccines

The BNT162b (Pfizer-BioNTech Inc., New York City, NY, USA) vaccine contains 30 μg of mRNA encoding the SARS-CoV-2 spike protein (S) (Polack et al., 2020). The mRNA- 1273 (Moderna Inc., Cambridge, MA, USA) contains 50 μg of mRNA encoding SARS-CoV-2 S protein, of which the half dose concentration has recently been recommended by the World Health Organization Strategic Advisory Group of Experts on Immunization (SAGE) as a booster dose for individuals aged 12 and older (Jackson et al., 2020, WHO, 2022). A dose of vaccine was administered intramuscularly.

### 2.3 Safety assessments

The enrolled participants self-reported reactogenicity using an electronic or paper questionnaire, starting on the day of vaccination and for 7 subsequent days (days 0–7). Local, systemic and adverse events (AE) were classified as mild, moderate and severe, as previously described (Kanokudom et al., 2022a).

### 2.4. Laboratory assessments

Serum samples were collected to determine binding antibody responses, including total immunoglobulin (Ig) anti-RBD of the SARS-CoV-2 spike protein (total RBD Ig) using the Elecsys SARS-CoV-2 S electrochemiluminescence immunoassay (ECLIA) according to the manufacturer”s instructions (Roche Diagnostics, Basel, Switzerland), the anti-RBD IgG using a Quant chemiluminescent microparticle immunoassay (CMIA) (Abbott Laboratories, Abbott Park, IL, USA), and anti-N IgG using CMIA (Abbott Diagnostics, Sligo, Ireland) as previously described (Kanokudom et al., 2022a). Neutralizing activity against the SARS-CoV-2 Spike protein RBD-HRP Omicron (BA.4/BA.5) (hereafter referred to as BA.4/5) (G339D, S371F, S373P, S375F, T376A, D405N, R408S, K417N, N440K, L452R, S477N, T478K, E484A, F486V, Q498R, N501Y, Y505H) (GenScript Biotech, Piscataway, NJ, USA) was analyzed using a surrogate virus neutralization test (sVNT) as previously described (Assawakosri et al., 2022b, Wanlapakorn N. et al., 2022b). Seropositivity of sVNT against Omicron (BA.4/5) was defined as ≥30% inhibition, respectively.

### 2.5. Statistical analysis

Baseline characteristics were reported as mean with standard deviation (SD) or median with interquartile range (IQR). Categorical age and sex analyses were performed using Pearson”s Chi-square test. The difference in incidence of an adverse event after the fourth dose with BNT162b2 or mRNA-1273 was analyzed using a risk difference (RD) with a 95% confidence interval (CI). Total RBD Ig and anti-RBD IgG were designated as geometric mean titers (GMT) with a 95% CI. The percentage inhibition using sVNT was presented as median with interquartile range (IQR) values. The geometric mean ratio (GMR) of total-RBD Ig and anti-RBD IgG was calculated using the Mann–Whitney *U* test. Significant differences between the two groups in antibody titers and percentage inhibition were compared using the Mann– Whitney *U* test. Non-parametric correlation between the GMT of total RBD Ig or anti-RBD IgG and the percent inhibition was calculated using Spearman”s rank correlation coefficient (Spearman”s ρ). The linear equation and coefficient of determination (r-square, R^2^) between the transformation of the logarithmic GMT of total-RBD Ig or anti-RBD IgG and percent inhibition was calculated based on a simple linear regression model. A *p*-value<0.05 was considered statistically significant.

## 3. Results

### 3.1 Demographic data and baseline characteristics

Between June and October 2022, a total of 292 healthy individuals from 12 different three-dose regimes (provided in **Figure 1**) were enrolled to receive a fourth booster dose of BNT162b2 or mRNA-1273. Sixty-five individuals were excluded and consisted of 24 subjects with pre-existing anti-N IgG, 8 subjects with anti-N IgG seroconversion from negative to positive, and 33 cases lost to follow-up. After exclusions, 227 individuals remained and were divided in two booster dose groups: BNT162b2 (n=109) and mRNA-1273 (n=118).

The demographics and characteristics of the eligible participants who received BNT162b2 or mRNA-1273, including sex, age, comorbidity, the time interval between the third and fourth dose, and the time interval between the fourth dose and blood collection (28±7 days) are shown in **Table 1**. The number of females (%) in the BNT162b2 and mRNA-1273 groups was 66.1% and 60.2%, respectively. The mean age of the BNT162b and mRNA-1273 groups was 48.8 and 48.2 years, respectively. There were no significant differences in terms of sex and age between the groups. The median interval between the third and fourth doses of BNT162b2 (191.0 days) was significantly shorter than that of the mRNA-1273 group (210.5 days).

**Table 1.** Characteristics of the individuals who received a fourth booster COVID-19 vaccine.

| Characteristic | BNT162b2 | mRNA-1273 |
| --- | --- | --- |
| Number | 109 | 118 |
| Sex, female n (%) | 72 (66.1) | 71 (60.2) |
| Age, years, mean [SD]<br>(min–max) | 48.8 [11.5]<br>(20.0–73.0) | 48.2 [11.8]<br>(23.0–74.0) |
| No comorbidity (%) | 68 (62.4) | 86 (72.9) |
| Underlying diseases (%) |  |  |
| Allergy | 5 (4.6) | 5 (4.2) |
| Dyslipidemia | 12 (11.0) | 11 (9.3) |
| Diabetes Mellitus | 4 (3.7) | 4 (3.4) |
| Hypertension | 11 (10.1) | 15 (12.7) |
| Thyroid | 2 (1.8) | 2 (1.7) |
| Others | 10 (9.2) | 4 (3.4) |
| Interval between the 3<sup>rd</sup> and 4<sup>th</sup> dose (days) |  |  |
| Median [IQR]<br>(min–max) | 191.0 [163.0–223.0]<br>(94.0–337.0) | 210.5 [177.0–224.3]<br>(117.0–371.0) |
| Interval between the 4<sup>th</sup> dose and blood collection (days) |  |  |
| Median [IQR]<br>(min–max) | 28.0 [28.0–29.0]<br>(22.0–32.0) | 28.0 [28.0–32.0]<br>(26.0–34.0) |

Abbreviation: IQR, interquartile range; SD, standard deviation.

### 3.2 Local and systemic adverse events resolved after receiving the mRNA-COVID-19 vaccine as the fourth booster dose

Profiles of local and systemic reactions within 7 days after vaccination with BNT162b2 and mRNA-1273 in each regimen included injection site pain, myalgia, and swelling (**Supplementary Figure S1A–L**). The safety profile of healthy participants who received a booster dose of BNT162b2 (n=139) or mRNA-1273 (n=150) was defined using a self-questionnaire. The most frequent adverse events were mild to moderate injection site pain (77.0% vs. 81.3%), followed by myalgia (63.3% vs. 62.0%), and swelling (39.6% vs. 54.0%) for the BNT162b2 and mRNA-1273 groups, respectively (**Figure 2A,B**). Furthermore, other adverse events observed in both groups were well tolerated and resolved in a few days. To compare the incidence between the two booster groups, swelling (RD: 14.4, 95%CI: 3.0–25.8), fever (RD: 4.0, 95%CI: 0.9–7.1) and headache (RD: 11.2, 95%CI: 0.4–22.1) were more frequent in the mRNA-1273 group than in the BNT162b2 group (**Figure 2C**).

**Figure 2.**
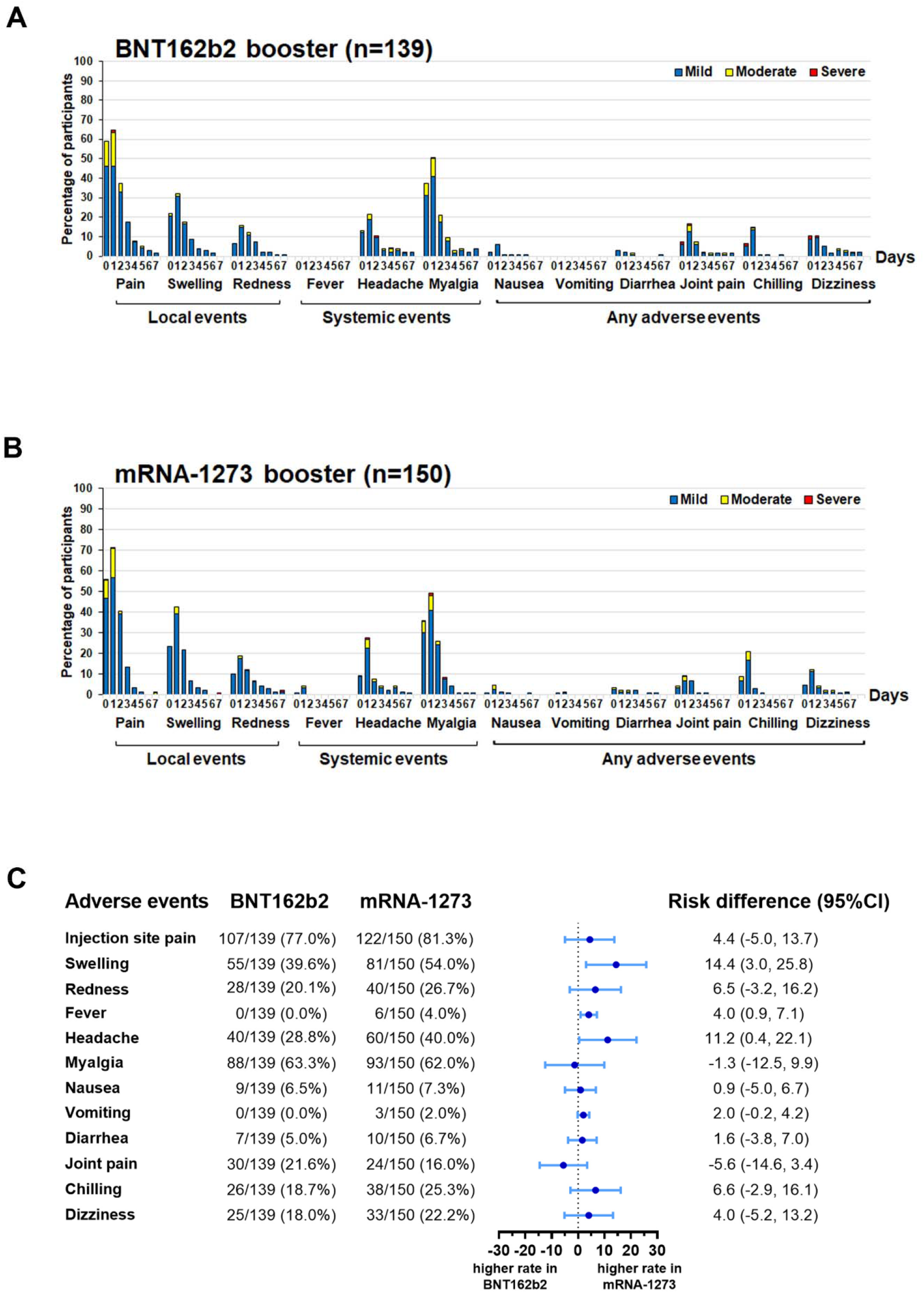
Overall reactogenicity of individuals who received a fourth dose of either BNT162b2 or mRNA-1273. The percentages of local, systemic, and any adverse events (AE) experienced within 7 days of receiving (A) BNT162b2 or (B) mRNA-1273. (C) Forest plots showing the incidences of each AE and the risk differences with 95% confidence intervals (95% CI).

### 3.3 Total-RBD Ig and anti-RBD IgG responses of eligible participants

A total of 227 individuals were seronegative anti-N IgG (no history of SARS-CoV-2 infection) and further examined for binding antibodies levels, including total RBD Ig and anti-RBD IgG. At baseline, the GMT of the total RBD Ig between 12 regimens ranged from 480.4 to 3112 U/mL. At 28 days after the fourth dose, all eligible individuals achieved a GMT of the total RBD Ig ranging from 11,992 to 32,003 U/mL (**Supplementary Figure S1A**). Furthermore, the GMT for the baseline anti-RBD IgG among all eligible individuals of the 12 regimens ranged from 77.9 to 492.2 BAU/mL. After the booster dose, the GMT of anti-RBD IgG for the 12 groups increased between 1562 and 4664 BAU/mL (**Supplementary Figure S1B)**. Although individuals had several three-dose schedules, the fourth dose generated a higher antibody titer specific for SARS-CoV-2 RBD for all regimens at 28 days.

To compare immunogenicity resulting from the fourth dose, sera samples were collected for the two groups according to the booster dose of BNT162b (n=109) and mRNA- 1273 (n=118). The baseline total RBD Ig GMT between the BNT162b2 and mRNA-1273 groups was similar (GMR: 0.93). After 28 days, the total RBD Ig GMT of the mRNA-1273 group was significantly higher than that of the BNT162b2 group (GMR: 1.17, *p<*0.05) **(Figure 3A)**. Similarly, the anti-RBD IgG response trends at baseline and after 28 days of receiving a fourth dose were consistent with the total RBD Ig response **(Figure 3B)**

**Figure 3.**
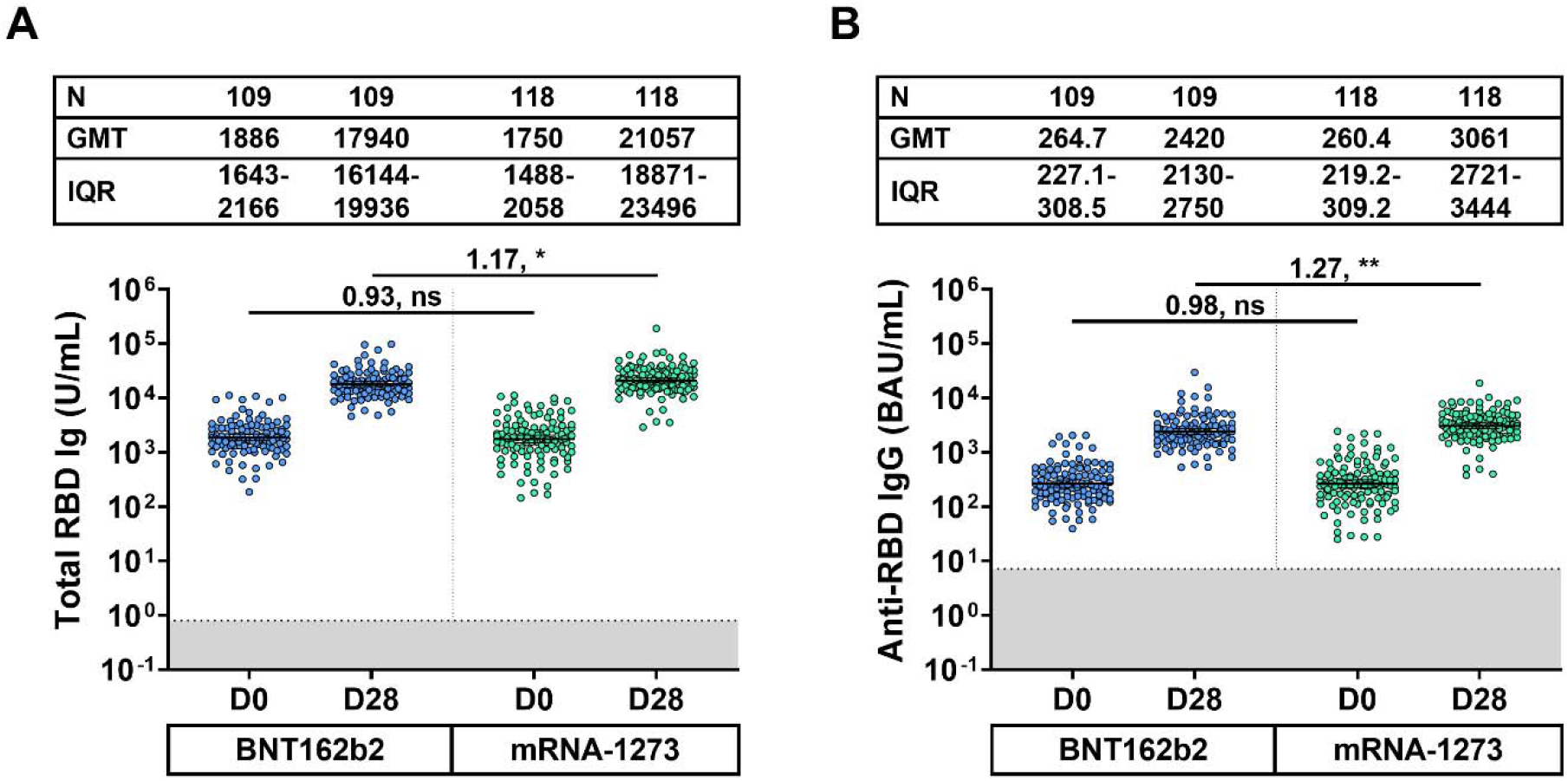
Binding antibody against the receptor binding domain of SARS-CoV-2 of the BNT162b2 and mRNA-1273 booster groups. (A) Total immunoglobulin (Ig) specific for the receptor binding domain (RBD) (Total RBD Ig) (U/mL) and (B) anti-RBD IgG (BAU/mL) of the BNT162b2 and mRNA-1273 booster groups were monitored at baseline (D0) and 28 days after vaccination (D28). The gray area indicates the seronegativity of total RBD Ig (<0.8 U/mL) or anti-RBG IgG (<7.1 BAU/mL) antibodies. Lines represent the geometric mean titer (GMT) with 95% confidence intervals (95% CI). A pairwise comparison shows geometric mean ratio (GMR) and statistical significance set at *p*<0.05 (*), *p*<0.001 (**), and no statistical significance (ns). Abbreviation: BAU, binding antibody unit; CI, confidence interval; D0, day 0; D28, day 28; GMT, geometric mean titer; GMR, geometric mean ratio; Ig, immunoglobulin; ns, no statistical significance.

### 3.4 Neutralizing activity against Omicron BA.4/5 using sVNT_50_

A subset of participants who received a fourth dose of BNT162b2 (n=67) and mRNA-1273 (n=93) was randomly selected for testing neutralizing activities against SARS-CoV-2 Omicron BA.4/5 using sVNT_50_. At baseline, the seronegativity (at threshold<30%) against Omicron BA.4/5 was 19/67 (28.4%) for BNT162b2 and 33/93 (35.5%) for the mRNA- 1273 groups. The median baseline neutralizing activity was 41.8% (IQR: 26.2–49.8%) for BNT162b2 and 35.9% (IQR: 22.0–50.2%) for the mRNA-1273 groups (**Figure 4**).

**Figure 4.**
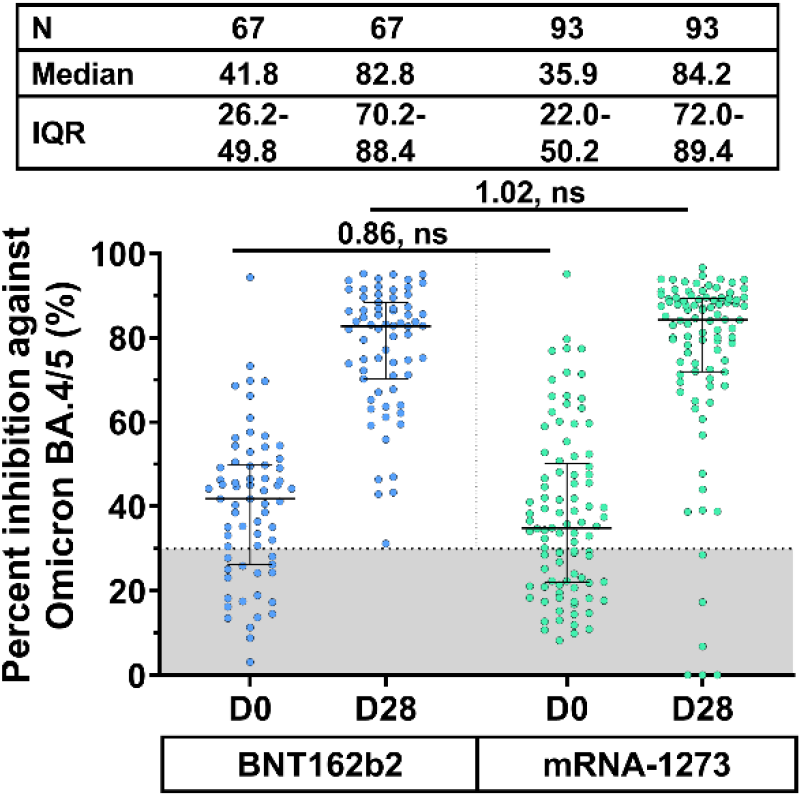
Neutralizing activity of participants against the SARS-CoV-2 Omicron BA.4/5 at 28 days after the fourth dose. Sera were obtained from a subset of individuals at baseline (D0) and at 28 days (D28) after receiving a booster dose of BNT162b2 and mRNA-1273. The gray area indicates the neutralizing activity of the SARS-CoV-2 Omicron BA.4/5 (<30%). Lines represent the medians (%) with interquartile range (IQR) values. A pairwise comparison displays the median ratio and the lack of statistical significance (ns). Abbreviation: D0, day 0; D28, day 28; GMT, geometric mean titer; GMR, geometric mean ratio; IQR; interquartile range; ns, no statistical significance.

The neutralizing activity of boosted individuals receiving BNT162b2 and mRNA- 1273 was significantly restored to 82.8% (*p<*0.001) and 84.2% (*p<*0.001) at 28 days, respectively. Furthermore, the median percentage inhibition against Omicron BA.4/5 between the BNT162b2 and mRNA-1273 groups was comparable to baselines (median ratio: 0.86) and after receiving a booster dose (median ratio: 1.02) (**Figure 4**).

### 3.5 Correlations between binding antibody and neutralizing activity against Omicron BA.4/5

To substantially assess the correlation between binding antibody and neutralizing activity against Omicron BA.4/5, a total of 320 sera were randomly selected from individual samples before (day 0) and after (day 28) receiving BNT162b2 or mRNA-1273. Quantitative ELISA levels of total RBD Ig and anti-RBD IgG, and *in vitro* ACE2-RBD binding neutralizing activity, which measured sVNT_50_, were further examined to determine the correlation between techniques. The percent inhibition for the Omicron BA.4/5 ranged from 0.0% (undetectable) to 99.6% compared to total RBD-Ig and anti-RBD IgG, respectively. The non-parametric correlation indicated that the percent inhibition against the Omicron BA.4/5 subvariant was strongly correlated with total RBD Ig (Spearman”s ρ: 0.83, *p*<0.001). A linear regression equation showing the percent inhibition against Omicron BA.4/5 based on log_10_(total RBD Ig) is presented in **Supplementary Figure 3A** (R^2^: 0.70). The correlation between the percent inhibition against the Omicron BA.4/5 and anti-RBD IgG showed a similar trend as total-RBD Ig (Spearman”s ρ: 0.82, *p*<0.001 and R^2^: 0.68) (**Supplementary Figure 3B**).

## 4. Discussion

We compared the reactogenicity of BNT162b2 and half-dose mRNA-1273 resulting from the administration of a fourth booster dose in healthy adults who had previously received different vaccine schedules. These findings revealed that common side effects were injection site pain, myalgia, headache, swelling, and redness at the injection site, which was in agreement with previous studies evaluating the application of mRNA vaccines as the third (Kanokudom et al., 2022a, Munro et al., 2021) and the fourth (Munro et al., 2022) doses.

This study evaluated the binding antibody targeting the SARS-CoV-2 spike protein in terms of total RBD Ig and anti-RBD IgG. For individuals who had received the third dose for approximately 3–12 months (min–max: 94–371 days), binding antibody levels were notably low. The present study showed that a fourth dose with the mRNA vaccine could restore the total RBD Ig titer from 11,992 to 32,003 U/mL among all regimens. Comparison of the two boosted groups showed that half-dose mRNA-1273 (50 μg) could induce a slightly higher response than in those immunized with BNT162b2 (30 μg). This evidence was possibly due to a dose-dependent response of active RNA molecules, as demonstrated previously (Kanokudom et al., 2022a). The GMT of total RBD Ig among 12 groups was varied due to abundant factors consisting of time intervals between the previous dose and the last dose, the vaccination dose, and the primary regimens. Additionally, in agreement with a previous study by Wanlapakorn et al. (2022), the present study indicated that individuals who were primed with an inactivated vaccine followed by an mRNA vaccine showed stronger immunogenicity compared to individuals who were primed with a viral vector or heterologous inactivated/viral vector vaccination schedule In the present study, the boosted neutralizing activity against the variant Omicron BA.4/5 after the fourth dose with the BNT162b2 and mRNA-1273 was observed on day 28. Our finding was in agreement with that of a previous study showing that there was no significant difference between BNT162b2 and mRNA-1273 in neutralizing activity against the Omicron variant (Wan et al., 2022). A fourth dose of BNT162b2 may increase the effectiveness of the vaccine against severe diseases compared to three doses (Magen et al., 2022). The fourth dose of BNT162b2 and mRNA-1273 subsequent to any three-dose regimen used in this study was expected to achieve a similar response to severe illness.

Our study evaluated the correlation between anti-RBD antibodies determined by two routine quantitative ELISA methods (expressed in total RBD Ig or anti-RBD IgG) and neutralizing activity determined by sVNT (expressed in percent inhibition of ACE2-RBD binding). Although two routine ELISA kits measured antibody titers against Wuhan-RBD and sVNT measured neutralizing antibody levels against Omicron BA.4/5, a strong correlation between the routine ELISA findings and sVNT was observed. Furthermore, our previous study evaluating different three-dose vaccinations showed a correlation between sVNT and the gold standard live virus neutralization test also called the foci-reduction neutralization test (FRNT_50_) against Omicron BA.1 and BA.2 (Spearman”s ρ: 0.77 and 0.64, respectively) (Suntronwong et al., 2022a). These data might suggest that the commercial sVNT assay was a suitable alternative to the conventional neutralization test.

The current findings demonstrated that boosted individuals with either mRNA vaccine could achieve a high level of binding antibody and neutralizing activity regardless of the primary three-dose vaccine regimens. Our findings were consistent with the VE study that showed that four-doses increased VE against Omicron infection compared to the three-doses or less (Intawong et al., 2022). In summary, these findings provide evidence for recommending the use of the mRNA vaccine as a fourth dose in healthy adults who completed three doses of mix-and-match as primary regimens.

This study was subject to certain limitations. It was challenging to find CIVID-19 naïve participants that had avoided multiple COVID-19 waves. Approximately 20% of the participants were excluded due to a suspected history of COVID-19 or were lost follow-up.

LongLJterm immunity and durability of immune responses after a fourth booster dose should be further determined. Additional studies on VE against infection and disease severity should be conducted to verify the findings.

## 5. Conclusions

This study found no significant difference in neutralizing activities against Omicron BA.4/5 among sera of individuals receiving either the BNT162b2 or mRNA-1273 vaccine as a fourth dose. The two routine ELISA methods, used to test total RBD Ig and anti-RBD IgG, could quantitatively predict neutralizing activity against Omicron BA.4/5. Overall, this study suggests that BNT162b2 and mRNA-1273 could be used as booster doses after different three-dose regimens.

## Data Availability

All data produced in the present study are available upon reasonable request to the authors

## Acknowledgments

We would like to thank all Center of Excellence in Clinical Virology personnel and all participants for contributing to and supporting this project. This research was financially supported by the Health Systems Research Institute (HSRI), National Research Council of Thailand (NRCT), MK restaurant Group Aunt Thongkum Foundation, BJC Big C Foundation, the Center of Excellence in Clinical Virology, Chulalongkorn University, and King Chulalongkorn Memorial Hospital, and was partially supported by the Second Century Fund (C2F) of Sitthichai Kanokudom, Chulalongkorn University.

## Author Contributions

Conceptualization, S.K. (Sitthichai Kanokudom), P.N., S.H., and Y.P.; data curation, S.K. (Sitthichai Kanokudom), R.Y., N.S. (Nungruthai Suntronwong), S.A., D.S., T.T. (Thaksaporn Thatsanatorn), N.S. (Natthinee Sudhinaraset) and N.W.; formal analysis, S.K. (Sitthichai Kanokudom); methodology, S.K. (Sitthichai Kanokudom), J.C., R.A., N.K., P.V., S.K. (Sirapa Klinfueng) and T.T (Thanunrat Thongmee); project administration, Y.P.; writing— original draft, S.K. (Sitthichai Kanokudom); writing—review and editing, S.K. (Sitthichai Kanokudom), S.H. and Y.P. All authors have read and agreed to the published version of the manuscript.

## Institutional Review Board Statement

The study protocol was approved by the Institutional Review Board (IRB), Faculty of Medicine, Chulalongkorn University (IRB number 223/65).

## Informed Consent Statement

Informed consent was obtained before participant enrollment. The study was conducted according to the Declaration of Helsinki and the Good Clinical Practice Guidelines (ICH-GCP) principles.

## Data Availability Statement

The datasets generated and analyzed during the current study are available from the corresponding author upon reasonable request.

## Supplementary materials

**Figure S1. Solicited local, systemic, and adverse events of enrolled participants in different regimens**. (A) SSAP, (B) SSAM, (C) SSPP, (D) SSMM, (E) AAAP, (F) AAAM, (G) AAPP, (H) AAMM, (I) SAAP, (J) SAAM, (K) SAPP, and (L) SAMM groups and the proportion of mild, moderate and severe adverse event 7 days after a booster dose for each group.

Abbreviations: A, AZD1222; M, mRNA-1273; P, BNT162b2; S, inactivated vaccine BBIBP-CorV or CoronaVac.

**Figure S2. Binding antibody titers against the receptor binding domain of SARS-CoV-2 of the 12 different four-dose regimens**. (A) Total immunoglobulin (Ig) specific for receptor binding domain (RBD) (Total RBD Ig) (U/mL) and (B) anti-RBD IgG (BAU/mL) were assessed for 12 different vaccine regimens on days 0 and 28 ± 7. The X-axis indicates the code of participant regimens, followed by the day of collection (0 and 28) The gray area indicates the seronegativity of total RBD Ig (<0.8 U/mL) or anti-RBG IgG (<7.1 BAU/mL). Each column shows a scatter plot with a geometric mean titer (GMT). The lines represent GMT with 95% confidence intervals (95% CI). Parentheses indicate the number of individuals in each regimen. Abbreviations: A, AZD1222; BAU, binding antibody unit; M, mRNA-1273; P, BNT162b2; S, inactivated vaccine BBIBP-CorV or CoronaVac.

**Figure S3. Predicted linear correlation between the binding antibody and neutralizing activity against Omicron BA.4/5 using the sVNT**_**50**_. A total of 320 sera (at day 0 (n=160) and day 28 (n=160)) from individuals who received a fourth dose of BNT162b2 and mRNA-1273 were tested for total RBD Ig and anti-RBD IgG levels and neutralizing activity against Omicron BA.4/5 using sVNT_50_. Linear regressions (solid line) showing the relationship between percent inhibition against Omicron BA.4/5 and logarithm (log_10_) (Total RBD Ig) (A) or log_10_ (Anti-RBD IgG) (B). The linear equation and the r-square (R^2^) were calculated according to a simple linear regression model between the transformed log_10_ of the binding antibody and the percentage of inhibition. The dotted line indicates the seronegativity of neutralizing activity of SARS-CoV-2 Omicron BA.4/5 (<30%).

Abbreviations: BAU, binding antibody unit; Ig, immunoglobulin.

