## Supplementary material for "The fourth dose of mRNA COVID-19 vaccine following 12 different three-dose regimens: Safety and immunogenicity to Omicron BA.4/BA.5": Supplementary Materials.docx

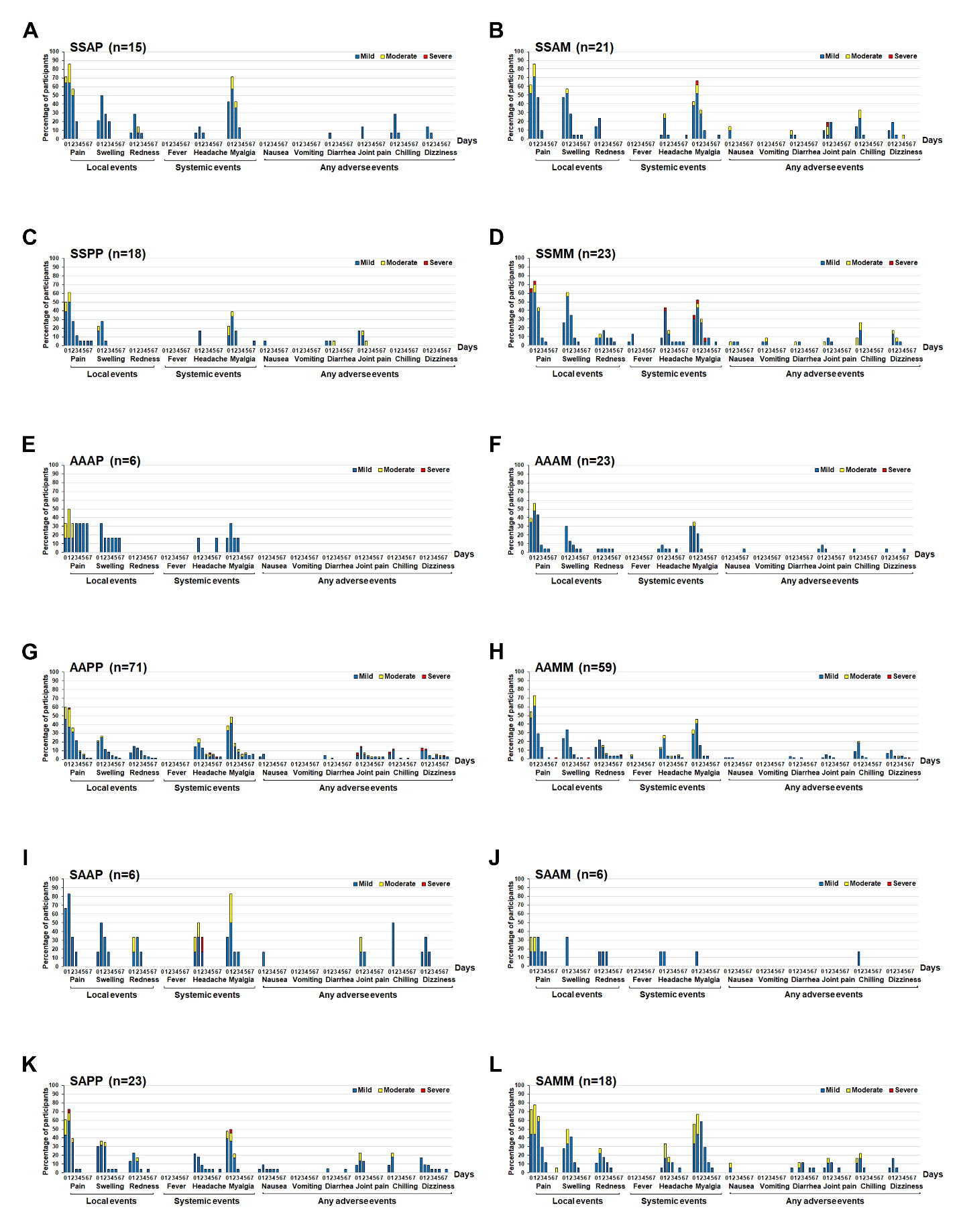

**Figure S1. Solicited local, systemic, and adverse events of enrolled participants in different regimens.** (A) SSAP, (B) SSAM, (C) SSPP, (D) SSMM, (E) AAAP, (F) AAAM, (G) AAPP, (H) AAMM, (I) SAAP, (J) SAAM, (K) SAPP, and (L) SAMM groups and the proportion of mild, moderate and severe adverse event 7 days after a booster dose for each group.

Abbreviations: A, AZD1222; M, mRNA-1273; P, BNT162b2; S, inactivated vaccine BBIBP-CorV or CoronaVac.

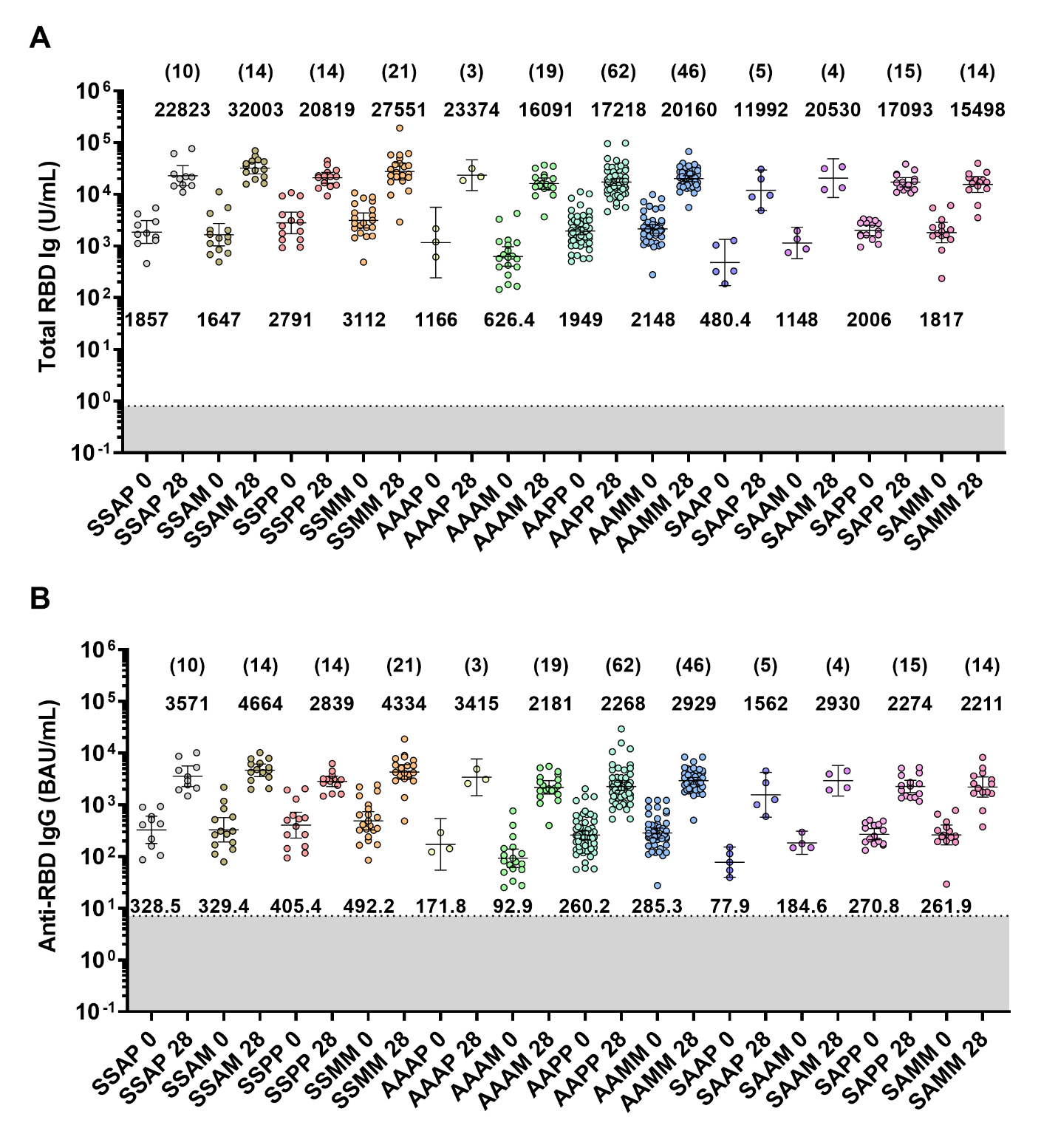

**Figure S2. Binding antibody titers against the receptor binding domain of SARS-CoV-2 of the 12 different four-dose regimens.** (A) Total immunoglobulin (Ig) specific for receptor binding domain (RBD) (Total RBD Ig) (U/mL) and (B) anti-RBD IgG (BAU/mL) were assessed for 12 different vaccine regimens on days 0 and 28 ± 7. The X-axis indicates the code of participant regimens, followed by the day of collection (0 and 28) The gray area indicates the seronegativity of total RBD Ig (<0.8 U/mL) or anti-RBG IgG (<7.1 BAU/mL). Each column shows a scatter plot with a geometric mean titer (GMT). The lines represent GMT with 95% confidence intervals (95% CI). Parentheses indicate the number of individuals in each regimen. Abbreviations: A, AZD1222; BAU, binding antibody unit; M, mRNA-1273; P, BNT162b2; S, inactivated vaccine BBIBP-CorV or CoronaVac.

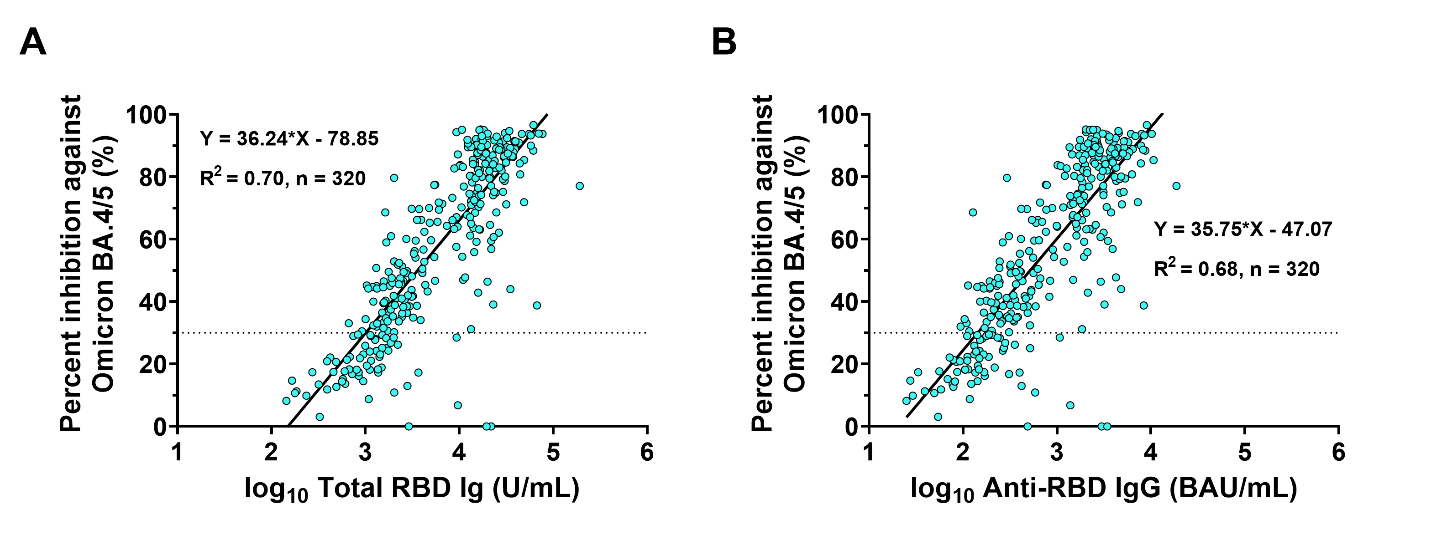

**Figure S3. Predicted linear correlation between the binding antibody and neutralizing activity against Omicron BA.4/5 using the sVNT_50_.** A total of 320 sera (at day 0 (n=160) and day 28 (n=160)) from individuals who received a fourth dose of BNT162b2 and mRNA-1273 were tested for total RBD Ig and anti-RBD IgG levels and neutralizing activity against Omicron BA.4/5 using sVNT_50_. Linear regressions (solid line) showing the relationship between percent inhibition against Omicron BA.4/5 and logarithm (log_10_) (Total RBD Ig) (A) or log_10_ (Anti-RBD IgG) (B). The linear equation and the r-square (R^2^) were calculated according to a simple linear regression model between the transformed log_10_ of the binding antibody and the percentage of inhibition. The dotted line indicates the seronegativity of neutralizing activity of SARS-CoV-2 Omicron BA.4/5 (<30%).

Abbreviations: BAU, binding antibody unit; Ig, immunoglobulin.
